# Biological variation estimates of Alzheimer’s disease plasma biomarkers in healthy individuals

**DOI:** 10.1101/2023.08.09.23293841

**Authors:** Wagner S. Brum, Nicholas J. Ashton, Joel Simrén, Guiglielmo di Molfetta, Thomas K. Karikari, Andrea L. Benedet, Eduardo R. Zimmer, Juan Lantero-Rodriguez, Laia Montoliu-Gaya, Andreas Jeromin, Aasne K. Aarsand, William A. Bartlett, Pilar Fernández Calle, Abdurrahman Coşkun, Jorge Díaz–Garzón, Niels Jonker, Henrik Zetterberg, Sverre Sandberg, Anna Carobene, Kaj Blennow, the European Federation of Clinical Chemistry and Laboratory Medicine Working Group on Biological Variation

**Affiliations:** Department of Psychiatry and Neurochemistry, Institute of Neuroscience and Physiology, The Sahlgrenska Academy at the University of Gothenburg, Mölndal, Sweden; Graduate Program in Biological Sciences: Biochemistry, Universidade Federal do Rio Grande do Sul (UFRGS), Porto Alegre, Brazil; King’s College London, Institute of Psychiatry, Psychology and Neuroscience Maurice Wohl Institute Clinical Neuroscience Institute London UK; NIHR Biomedical Research Centre for Mental Health and Biomedical Research Unit for Dementia at South London and Maudsley NHS Foundation London UK; Centre for Age-Related Medicine, Stavanger University Hospital, Stavanger, Norway; Clinical Neurochemistry Laboratory, Sahlgrenska University Hospital, Mölndal; Department of Psychiatry, University of Pittsburgh, Pittsburgh, PA, USA; Department of Pharmacology, Universidade Federal do Rio Grande do Sul (UFRGS); Porto Alegre, RS, Brazil; Graduate Program in Biological Sciences: Pharmacology, Universidade Federal do Rio Grande do Sul (UFRGS), Porto Alegre, Brazil; McGill Centre for Studies in Aging, McGill University, Montreal, QC, Canada; ALZpath. Inc, Carlsbad, CA 92008, USA; The Norwegian Organization for Quality Improvement of Laboratory Examinations (NOKLUS), Haraldsplass Deaconess Hospital, Bergen, Norway; School of Science and Engineering, University of Dundee Dundee – UK; Department of Laboratory Medicine, La Paz University Hospital, Madrid, Spain; Acibadem Mehmet Ali Aydınlar University, School of Medicine, Department of Medical Biochemistry, Atasehir, Istanbul, Turkey; Certe, Wilhelmina Ziekenhuis Assen, Assen, The Netherlands; Department of Neurodegenerative Disease, UCL Institute of Neurology, London, UK; UK Dementia Research Institute at UCL, London, UK; Hong Kong Center for Neurodegenerative Diseases, Clear Water Bay, Hong Kong, China; Wisconsin Alzheimer’s Disease Research Center, School of Medicine and Public Health, University of Wisconsin-Madison, Madison, WI 53792, USA; Department of Global Health and Primary Care, Faculty of Medicine, University of Bergen, Bergen, Norway; Laboratory Medicine, IRCCS San Raffaele Scientific Institute, Milano, Italy

**Author notes:** **Corresponding authors: Kaj Blennow**, Department of Psychiatry and Neurochemistry, Institute of Neuroscience and Physiology, University of Gothenburg, Mölndal, Sweden;, **Anna Carobene,** Laboratory Medicine, IRCCS San Raffaele Scientific Institute, Milan, Italy. Contributed equally as senior authors (Anna Carobene, Kaj Blennow).

**Keywords:** biological variation, analytical variation, plasma biomarkers, p-tau, amyloid, glial fibrillary acidic protein, neurofilament light, reference change values

## Abstract

**Introduction:** Blood biomarkers have proven useful in Alzheimer’s disease (AD), but little is known about their biological variation (BV), which plays a crucial role in the interpretation of individual patient data.

**Methods:** We measured plasma amyloid-β (Aβ42, Aβ40), phosphorylated tau (p-tau181, p-tau217, p-tau231), glial fibrillary acidic protein (GFAP), and neurofilament light chain (NfL) in fasting plasma samples collected weekly over 10 weeks from 20 participants aged 40-60y from the European Biological Variation Study. We determined within- (CV_I_) and between-subject (CV_G_) BV, analytical variation (CV_A_) and reference change values (RCV).

**Results:** Biomarkers presented considerable variability in CV_I_ and CV_G_. Aβ42/Aβ40 had the lowest CV_I_ (∼3%) and p-tau181 the highest (∼16%), while the others ranged from 6-10%. Most RCVs ranged from 20-30% (decrease) and 25-40% (increase).

**Interpretation:** We provide BV estimates for AD plasma biomarkers, which can potentially refine their clinical and research interpretation. RCVs might be useful for detecting significant changes between serial measurements when monitoring early disease progression or interventions.

**Highlights:**

· Plasma Aβ42/Aβ40 presents the lowest between- and within-subject biological variation, but also changes the least in AD patients vs controls.
· Plasma p-tau variants significantly vary in their within-subject biological variation, but their substantial fold-changes in AD likely limits the impact of their variability.
· Plasma NfL and GFAP demonstrate high between-subject variation, the impact of which will depend on clinical context.
· Reference change values can potentially be useful in monitoring early disease progression and the safety/efficacy of interventions on an individual-level.
· Serial sampling revealed that unexpectedly high values in heathy invidivuals can be observed, which urges caution when interpreting AD plasma biomarkers based on a single test result.

**Research in Context:** *Systematic Review:* We reviewed PubMed for articles and conference abstracts that evaluated the biological variation (BV) of novel Alzheimer’s disease (AD) blood biomarkers. Two previous studies had reported BV estimates for serum glial fibrillary acidic protein (GFAP) and neurofilament light chain (NfL). Thus, we aimed to provide the first robust BV estimates for plasma amyloid-β (Aβ) and phosphorylated tau (p-tau) biomarkers, and also for plasma GFAP and NfL in in the same population.

*Interpretation:* Plasma biomarkers of key pathological features of AD demonstrate heterogeneity in their within- and between-subject variation. Plasma Aβ42/Aβ40 generally shows lower variability but also changes very modestly in AD patients vs controls. While plasma p-tau variants demonstrate higher variability, its clinical impact is likely limited due to large fold-increases in AD. Plasma NfL and GFAP had the largest between-subject variability, which may impact upon their application in certain contexts. Most research on blood biomarkers so far has been done using either single measurements or repeated measurements over longer (e.g., yearly) time frames; the weekly serial sampling in our study revealed that unexpected outlier values may occur, urging caution in clinical and research interpretation.

*Future directions:* Future studies should evaluate the potential clinical impact of the application of BV knowledge upon clinical and research interpretation of AD plasma biomarkers, especially in disease monitoring and in the evaluation of safety and efficacy of novel therapeutic interventions.

## 1. Introduction

Novel technologies to measure brain pathophysiological processes in the blood have revolutionized the Alzheimer’s disease (AD) research landscape.^1,2^ Established and highly accurate methods for tracking such processes face barriers to their large-scale implementation, such as the high costs, radiation exposure and limited availability of positron emission tomography (PET) scans, as well as the relative invasiveness of lumbar punctures, required for measuring AD biomarkers in the cerebrospinal fluid (CSF).^3^ Blood-based AD biomarkers have demonstrated great promise so far, and are particularly promising for scalable implementation due to their minimally invasive and cost-effective nature.^1,2^

Among blood-based biomarkers so far investigated, plasma phosphorylated tau (p-tau) variants, such as p-tau181, p-tau231 and p-tau217, have demonstrated the greatest potential to identify AD-specific processes, showing high accuracy for identifying neuropathological or biomarker-confirmed AD and predicting cognitive decline.^4–8^ While p-tau231 may be more sensitive to incipient amyloid-β (Aβ) pathology, plasma p-tau217 seems the most well-suited for clinical implementation, presenting the highest fold-increases in cognitively impaired patients with AD-type pathology, and it can dynamically track longitudinal AD clinical progression.^4,6,7,9–11^ Plasma Aβ, in the form of the Aβ42/Aβ40 ratio, has also shown good performance in detecting Aβ pathology, but its modest fold-change (reduced by 8-14% in AD compared with Aβ-negative controls, when in the CSF it is reduced by >50%)^12,13^ makes it more vulnerable to analytical fluctuations normally observed in a day-to-day clinical chemistry routine.^14–16^ Plasma levels of glial fibrillary acidic protein (GFAP), a cytoskeletal protein highly expressed in reactive astrocytes^17^, have been positively associated with early Aβ pathology.^18–21^ Neurofilament light (NfL), a marker for axonal damage, has gained increasingly clinical significance with robust evidence for its diagnostic and prognostic utility in a wide range of neurodegenerative diseases (AD, frontotemporal dementia, atypical parkinsonian disorders) and in acute neurological conditions, such as stroke and traumatic brain injury.^22–26^ Furthermore, all of these biomarker candidates have been evaluated as potential surrogate endpoints disease-modifying clinical trials in AD, with a recent example being reductions in plasma p-tau217 as early as after 12 weeks of treatment with a promising anti-Aβ monoclonal antibody, donanemab.^27^

Nevertheless, several research questions must be addressed before the large-scale implementation of blood-based AD biomarkers.^28^ While most studies focused on their diagnostic and prognostic properties, little is known about their biological variation (BV) – a foundational concept in clinical chemistry, crucial to ensure the safe implementation of diagnostic markers and to minimize misclassification risks in laboratory medicine.^29^ BV refers to the variation observed in clinical laboratory measurements determined by patients’ physiology, and a strict guideline-defined methodology must be followed by BV studies to ensure robust results.^30,31^ Such studies require the serial, tightly controlled collection of samples from healthy individuals with a regular sampling rate, and that analytes should be quantified, at least, in duplicate.^30,31^ The key BV components are the within-subject biological variation (CV_I_), which informs how much the concentration of a biomarker fluctuates around each individual’s homeostatic setpoint, and the between-subject biological variation (CV_G_), which informs on the variability between the homeostatic setpoints between different individuals. These parameters, alongside known assay-dependent analytical variation (CV_A_), can provide highly clinically useful information for biomarker implementation. These include the reference change value (RCV),^32,33^ which enumerates the change needed between consecutive measurements to exceed biological and analytical variation, the analytical performance specifications (APS) that clinical-grade assays should meet,^34,35^ and the index of individuality (II), which evaluates the utility of population based reference intervals^33,36^. Thus, high-quality BV data is needed in this rapidly developing area of AD diagnostics, in which specific biomarkers and assays are being considered for clinical implementation and therapeutic trial use.

Here, we aimed to determine BV estimates for plasma Aβ42, Aβ40, Aβ42/Aβ40, p-tau181, p-tau217, p-tau231, GFAP and NfL (and associated APS and RCVs) in healthy adults between 40-60 years from the European Biological Variation Study (EuBIVAS), led by the European Federation of Laboratory Medicine (EFLM) Working Group on Biological Variation.^37,38^ The EuBIVAS is a highly powered multi-center study that included weekly blood sampling over 10 weeks from presumably healthy volunteers from five European countries and that has delivered high-quality BV estimates for many measurands, widely used in diverse medical areas.^38–42^

## 2. Methods

### 2.1 Study participants and sample collection

In this study, we quantified biomarkers in plasma-citrate samples from a subset of 20 individuals aged between 40-60 years within the EuBIVAS,^37,38^ which originally enrolled 91 healthy volunteers (53 females; 38 males; ages 21-69 years), from six European laboratories located in five different countries (Italy, Norway, Spain, Turkey and the Netherlands). We chose to include in the current study those in the older EuBIVAS age range that had sufficient sample material for analyses. Information on the participants’ health status and lifestyle was collected with an enrollment questionnaire, and participants were screened at enrollment with a selection of laboratory tests to further confirm compatibility with inclusion criteria. Fasting blood samples were collected weekly over 10 consecutive weeks for each study participant (April-June 2015), always in the morning. At each center, samples were centrifuged at 3000 g for 10 min at room temperature within 1h of the blood draw, aliquoted and frozen rapidly by immersion in a bowl with methanol and dry ice, and sent to the coordinating center (San Raffaele Hospital in Milan, Italy), where they were stored at -80°C. In November 2021, the samples included in this study were sent to the Clinical Neurochemistry Laboratory (Sahlgrenska University Hospital, Gothenburg, Sweden), where the AD blood biomarkers were measured (April 2022, except p-tau217, analyzed December 2022). Further details regarding the inclusion/exclusion criteria, health status, and sample collection, processing, and storage protocol used in EuBIVAS have been previously reported.^37^

The protocol for EuBIVAS received approval from the Institutional Ethical Review Board of San Raffaele Hospital, in compliance with the World Medical Association Declaration of Helsinki, as well as the Ethical Board/Regional Ethics Committee for each participating center.

### 2.2 Biomarker quantification

Biomarker quantification was conducted using Single molecule array (Simoa) HD-X Analyzers from Quanterix (Billerica, MA/USA) at the Clinical Neurochemistry Laboratory of Sahlgrenska University Hospital in Sweden. A commercially available assay (Quanterix Neurology-4 Plex E) was used to simultaneously quantify for Aβ42, Aβ40 (and Aβ42/Aβ40, consequently), NfL, and GFAP.^27^ P-tau231 and p-tau181 were analyzed using Simoa assays developed at the University of Gothenburg, which have been validated as described elsewhere.^5,6^ To measure p-tau217, a novel commercially available assay from ALZpath (ALZpathDX, Carlsbad, CA/USA) was used.^43^ All samples from the same participant were analyzed in the same analytical run, and each sample was quantified in duplicate. Internal quality controls (iQC) at three different concentrations, for each measurand, were analyzed in duplicate in the beginning and end of each run. Before analysis, blood samples were thawed, vortexed, and centrifuged at 4000 × g for 10 minutes as suggested in recent studies.^44,45^

### 2.3 Statistical analysis

Our statistical analyses followed a series of well-established and guideline-defined steps for deriving BV data, as set out by the Biological Variation Data Critical Appraisal Checklist (BIVAC), a standard for the executing and reporting of BV studies.^31^ Outlier detection procedures were performed on three levels, including analytical (between replicates), within-subject (among 10 collections for CV_I_ calculation), as well as on the between-subject level (for CV_G_ calculation).^46–50^ For obtaining CV_I_ and CV_A_ estimates, we initially performed CV-transformation of the data where each person’s data is “normalized” by dividing it by that person’s mean value, so as to later perform the ANOVA on these CV-transformed values.^51^ After CV-transformation, we performed outlier identification and removal on the analytical levels (between replicates) by assessing the homogeneity of CV_A_ with the Bartlett test. In case of heterogeneity for the analytical component, we first excluded the replicate value of the measurement that most deviated from that participant’s mean. If the heterogeneity persisted, we then also excluded the second measurement result of the time-point showing abnormal analytical variation. After ensuring analytical homogeneity, we evaluated the presence of outliers on the within-individual variation level by assessing the homogeneity of the within-individual CV_I_ with the Cochran test. Then, we evaluated for each biomarker, whether the results were consistent with steady state (i.e. no trend for increase or decrease during study) by fitting a linear regression model with the mean blood drawing value (pooled mean of the duplicate concentration measurements of each participant) as the dependent variable, with blood drawing number (from 1-10) as the independent variable. Individuals were considered to be in a steady state if the 95% CI of the blood draw term (i.e., the slope) included 0.^52^ Finally, the CV_I_ was estimated with CV-ANOVA, the “Røraas method”, a validated and recommended ANOVA method for estimating CV_I_ and CV_A_.^51,53^ To calculate the between-subject biological variation (CV_G_), we first applied the Dixon-Q test to detect outliers in mean biomarker concentrations between subjects, and the Shapiro-Wilk test to verify the normality assumption on mean concentrations. If the latter tests detected a non-normal distribution, concentration data were natural log-transformed, prior to obtaining the CV_G_ by ANOVA.^46–50^ First, CV_I_ and CV_G_ estimates were calculated for the whole study population, and also secondarily separately for males and females, for all measurands. Confidence intervals (CIs) for BV estimates were calculated as previously described,^54^ and the lack of overlap of the 95% CI of estimates was used to indicate significant differences between subgroups.

Other relevant metrics were computed based on the above-mentioned BV estimates were calculated as follows. Desirable analytical performance specifications (APS) were calculated for imprecision (CV_APS_ = 0.5_x_CV_I_) and for bias [Bias_APS_ = 0.25_x_√(CV_I_^2^ + CV_G_^2^)]. The reference change value (RCV) was calculated at a 95% bidirectional alpha (z=1.65) as RCV = 100*(exp (±z_x_√2_x_σ)–1), where σ=√ln(σ^2^_CVi_ + σ^2^_CVi_), with σ^2^_CVi_ = ln(CVi^2^ + 1) and σ^2^_CVa_ = ln(CVa^2^ + 1). The index of individuality (II) was calculated as the ratio of CV_I_ and CV_G_ for each biomarker, and indicates whether population-based reference intervals can be useful for evaluating results.^33,36^ We also calculated the number of samples needed to be collected to estimate an individual’s homeostatic point (NHSP) with a “D” absolute percentage proximity to the individual’s true value with the equation n = (z_x_√(CV_I_^2^ + CV_A_^2^)/D)^2^, in which z=1.96, corresponding to a 95% alpha. NHSP was calculated based on 5, 10 and 20% deviations from the homeostatic setpoint. Metrics such as RCVs and APS were always derived based on CV_I_ and CV_G_ of all participants. All analyses were performed with R Statistical Software (version 4.2.1; www.r-project.com), and statistical significance was set as alpha=0.05.

## 3. Results

### 3.1 Participant characteristics

We included data analyzed from a total of 196 plasma samples, collected weekly over 10 weeks from 20 participants, with a mean number of 9.8 samples per participant. Key demographic information is described in Table 1. The age range of the included participants was 40-60 years, with a mean (SD) age of 46.4 years (6.20) for the whole study population. Half of the participants were female, and key demographic characteristics were generally similar between sexes. The study population came from 5 centers in 4 European countries (Italy [n=7], Netherlands [n=5], Norway [n=5], Spain [n=3]), and all participants were white/caucasian. Participants were healthy, with a mean (SD) body mass index of 23.3 kg/m^2^ (2.85 kg/m^2^), did not have hypertension, and the majority (55%) engaged in physical activity for more than 3 hours per week. Only one participant was a smoker (5%), and 11 reported consuming 1-2 units of alcohol per week.

**Table 1.** Demographic characteristics.

|  | Female<br>(n=10) | Male<br>(N=10) | Overall<br>(N=20) |
| --- | --- | --- | --- |
| Age, years, mean (SD) | 47.0 (5.98) | 45.8 (6.68) | 46.4 (6.20) |
| BMI, kg/m<sup>2</sup>, mean (SD) | 22.3 (2.50) | 24.2 (2.98) | 23.3 (2.85) |
| Hypertension, n (%) | 0 | 0 | 0 |
| Alcohol consumption, units/week, n (%) |  |  |  |
| 0 | 3 (30.0) | 3 (30.0) | 6 (30.0) |
| 1-2 | 6 (60.0) | 5 (50.0) | 11 (55.0) |
| ≥ 3 | 1 (10.0) | 2 (20.0) | 4 (15.0) |
| Smokers, n (%) | 1 (10) | 0 | 1 (5) |
| Physical exercise, n (%) | 8 (80) | 5 (50) | 13 (65) |
| No physical exercise | 2 (20) | 5 (50) | 7 (35) |
| < 3h per week | 1 (10) | 1 (10) | 2 (10) |
| ≥ 3 | 7 (70) | 4 (40) | 11 (55) |
| Study center, n (%) |  |  |  |
| Italy (Milan) | 2 (20) | 2 (20) | 4 (20) |
| Italy (Padua) | 3 (30) | 0 | 3 (15) |
| Netherlands | 3 (30) | 2 (20) | 5 (25) |
| Norway | 2 (20) | 3 (30) | 5 (25) |
| Spain | 0 | 3 (30) | 3 (15) |

The table summarises key demographic information for the included participants. Data are described as mean (SD) or n (%).

### 3.2 Homogeneity analyses and outliers

Table 2 displays results of the homogeneity analyses for outlier detection and the final number of results included for each of the biomarkers. All samples were always analyzed in duplicate, except in very few cases with insufficient volume left, resulting in a mean of 1.97 replicate quantification per sample per biomarker. When evaluating the analytical homogeneity with the Bartlett test, no outliers for the replicate measurement were identified for Aβ40, Aβ42/Aβ40, GFAP, and NfL, while a few replicates were excluded for Aβ42, P-tau181, P-tau217, and P-tau231. When assessing the variance homogeneity for within-subject variation, outlier time points were identified for all biomarkers, but no subject had to be fully excluded. For the total study population, the mean percentage of results identified as outliers at the homogeneity analyses was 3.56% (range, 1.0–6.8%), which left a mean of 369 results (range, 332–381) used per biomarker to estimate the CV_I_.

**Table 2.**
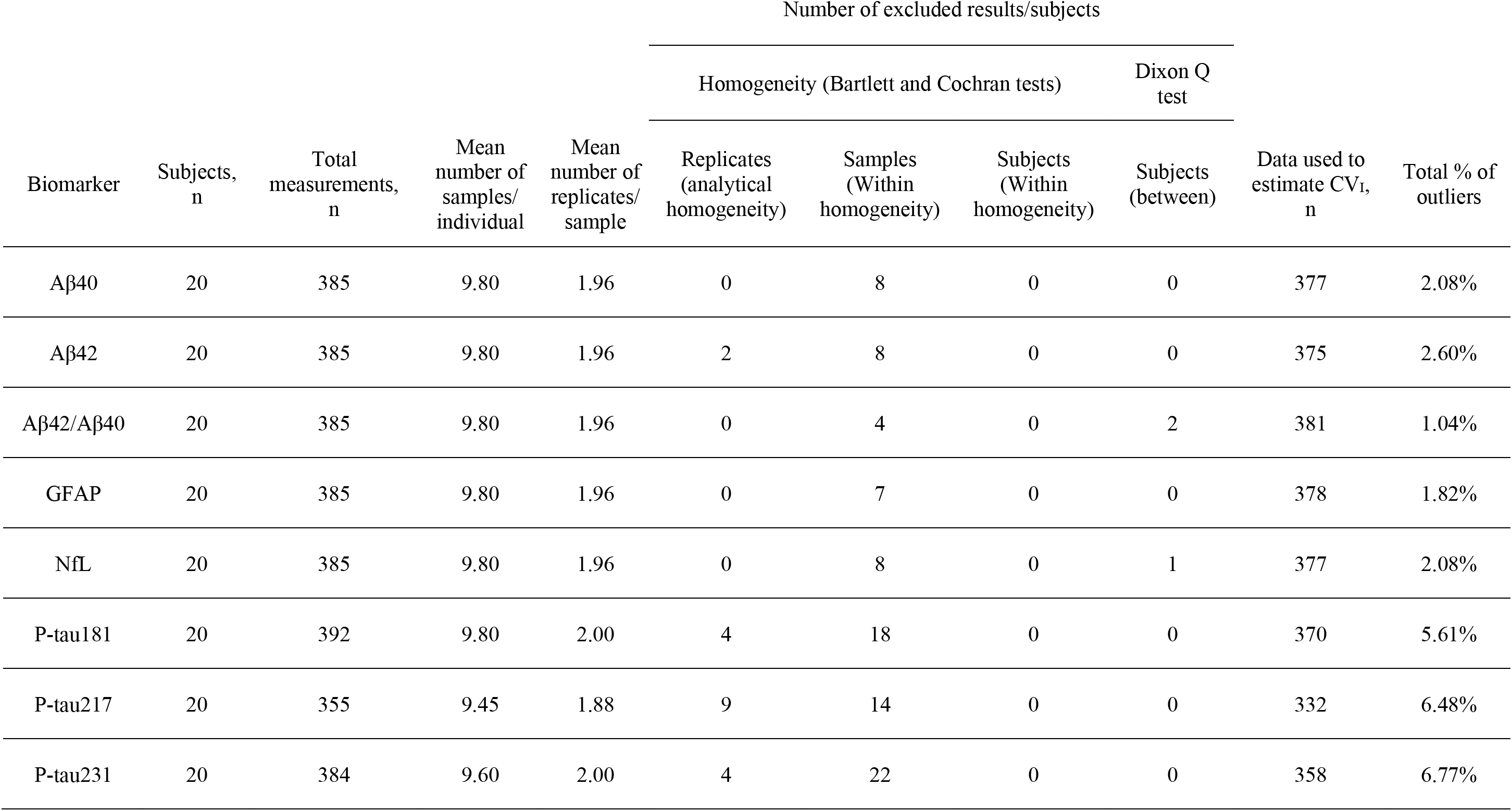
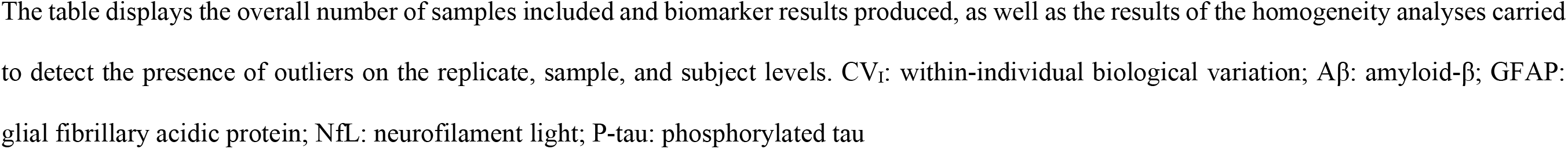
Homogeneity analyses and number of results included for calculation of biological variation estimates.

| Biomarker | Subjects,<br>n | Total<br>measurements,<br>n | Mean<br>number of<br>samples/<br>individual | Mean<br>number of<br>replicates/<br>sample | Number of excluded results/subjects |  |  |  | Data used to<br>estimate CV<sub>i</sub>,<br>n | Total % of<br>outliers |
| --- | --- | --- | --- | --- | --- | --- | --- | --- | --- | --- |
|  |  |  |  |  | Homogeneity (Bartlett and Cochran tests) |  |  | Dixon Q<br>test |  |  |
|  |  |  |  |  | Replicates<br>(analytical<br>homogeneity) | Samples<br>(Within<br>homogeneity) | Subjects<br>(Within<br>homogeneity) | Subjects<br>(between) |  |  |
| Aβ40 | 20 | 385 | 9.80 | 1.96 | 0 | 8 | 0 | 0 | 377 | 2.08% |
| Aβ42 | 20 | 385 | 9.80 | 1.96 | 2 | 8 | 0 | 0 | 375 | 2.60% |
| Aβ42/Aβ40 | 20 | 385 | 9.80 | 1.96 | 0 | 4 | 0 | 2 | 381 | 1.04% |
| GFAP | 20 | 385 | 9.80 | 1.96 | 0 | 7 | 0 | 0 | 378 | 1.82% |
| NfL | 20 | 385 | 9.80 | 1.96 | 0 | 8 | 0 | 1 | 377 | 2.08% |
| P-tau181 | 20 | 392 | 9.80 | 2.00 | 4 | 18 | 0 | 0 | 370 | 5.61% |
| P-tau217 | 20 | 355 | 9.45 | 1.88 | 9 | 14 | 0 | 0 | 332 | 6.48% |
| P-tau231 | 20 | 384 | 9.60 | 2.00 | 4 | 22 | 0 | 0 | 358 | 6.77% |

The table displays the overall number of samples included and biomarker results produced, as well as the results of the homogeneity analyses carried to detect the presence of outliers on the replicate, sample, and subject levels. CV<sub>I</sub>: within-individual biological variation; Aβ : amyloid-β ; GFAP: glial fibrillary acidic protein; NfL: neurofilament light; P-tau: phosphorylated tau

In Figure 1 the 10-week biological variation, in concentrations, of each plasma biomarker, stratified by sex and ordered by increasing age is displayed. In a separate outlier detection procedure before the CV_G_ estimation, the Dixon-Q test identified 1 outlier subject for NfL, and 2 outlier subjects for Aβ42/Aβ40 (indicated in Figure 1). No trend was identified for any of the included biomarkers in the overall study population or in male or female subgroups. No biomarker measurement for any analyte was below the lower limit of detection or the lower limit of quantification.

**Figure 1.**
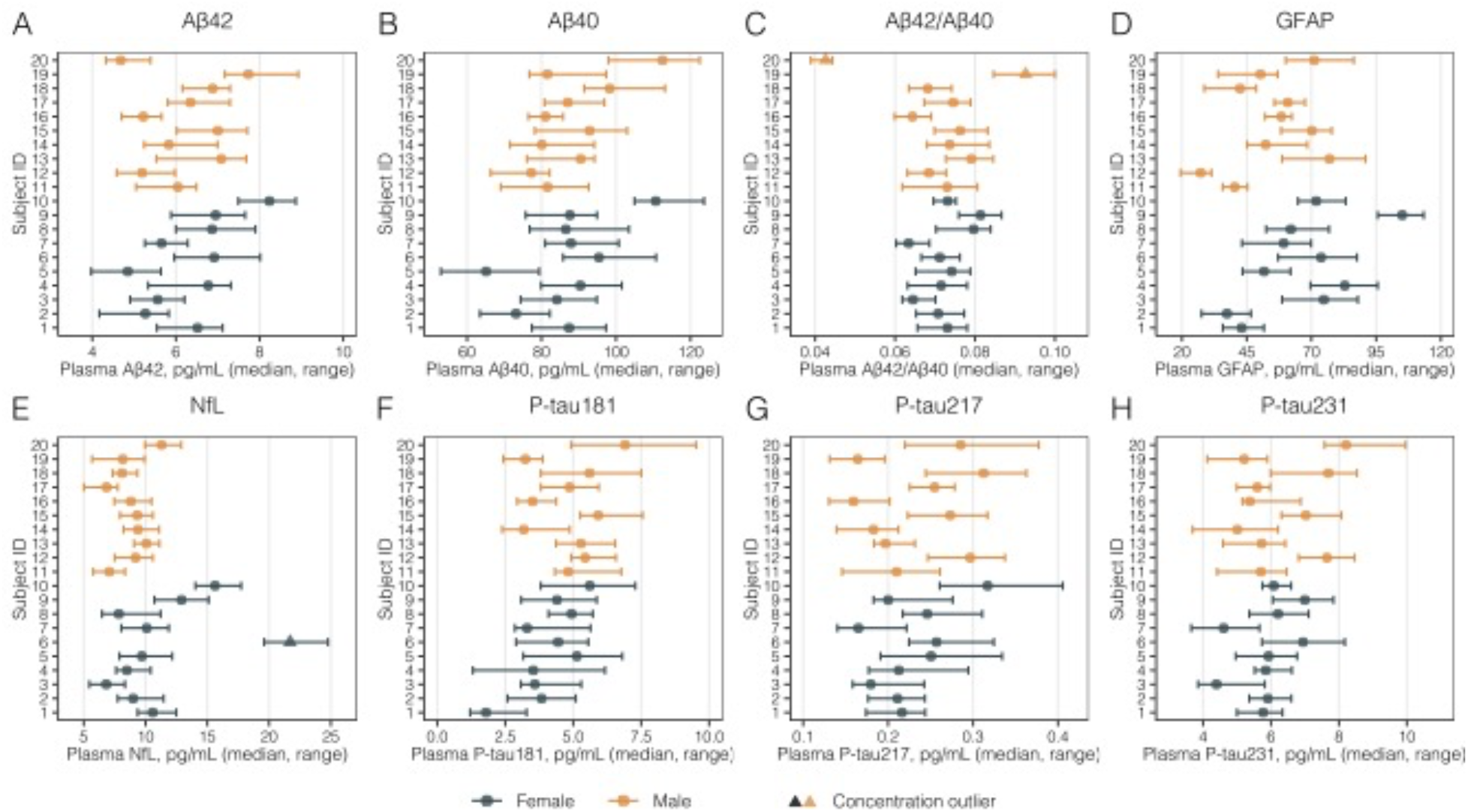
Participant-level plasma biomarker concentrations over 10 weeks. The figure displays median (dots) and range (errorbars) of biomarker concentrations over 10 weeks. Females are represented in dark green, and males in orange, and participants are shown with increasing age (subject 1 is the youngest female participant, and subject 20 the oldest male). Triangles represent the concentration outliers detected with the Dixon-Q test before the CV_G_ calculation. CV_G_: between-subject biological variation; Aβ: amyloid-β; GFAP: glial fibrillary acidic protein; NfL: neurofilament light; P-tau: phosphorylated tau.

### 3.3 Analytical performance (CV_A_)

The CV_A_ for each biomarker, which indicates the imprecision between duplicate measurements, and associated 95% CIs are graphically displayed in Figure 2A and numerically represented in Table 3. The CV_A_ ranged from around 3% for all Aβ biomarkers (Aβ42: 2.8%; Aβ40: 2.6%; Aβ42/Aβ40: 3.0%), to around 6% for GFAP (6.4%) and NfL (6.3%), and to approximately 5.5% for all p-tau biomarkers (p-tau181: 5.6%; p-tau217: 5.7%; p-tau231: 5.6%). Analytical variability of internal quality controls presented similar CVs to those estimated with CV-ANOVA, and no systematic trends in concentration change between-runs were observed by visual inspection.

**Figure 2.**
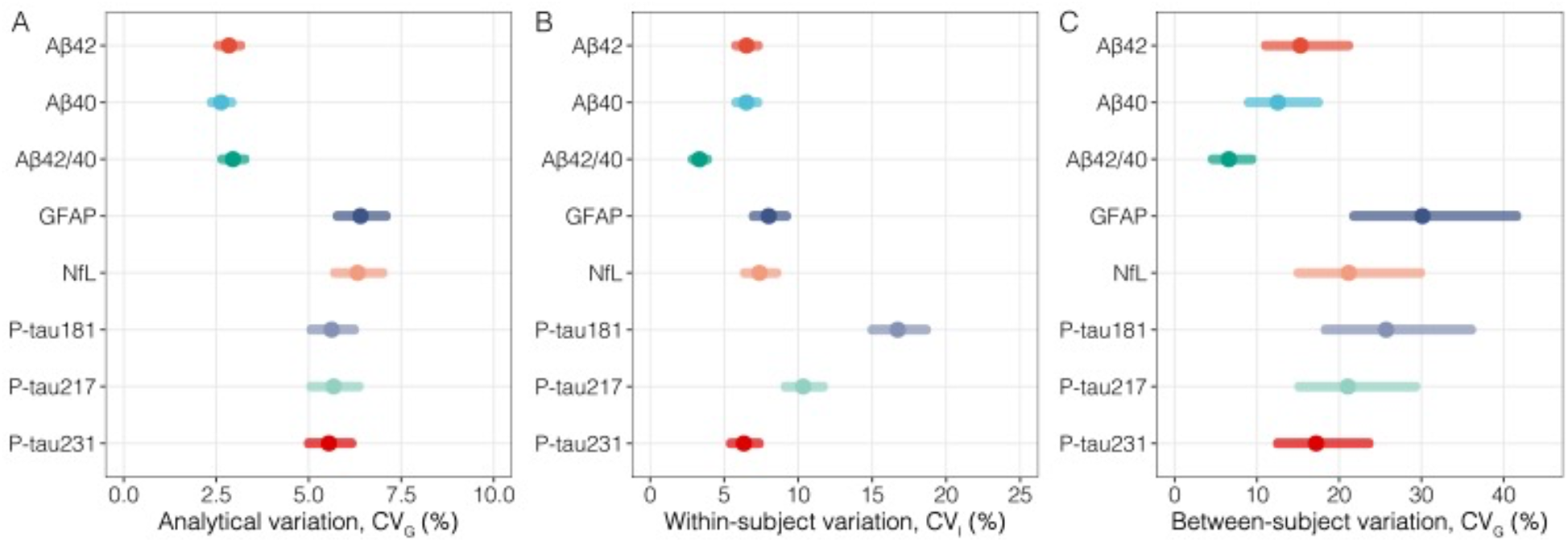
Biological variation estimates in the whole study population. The forest plot graphically summarises the biological variation estimates obtained in this study. (A) The left panel indicates analytical variation (CV_A_); (B) The middle panel displays estimates for within-individual biological variation; (C) The right panel indicates between-individual biological variation. CV_I_: within-subject biological variation; CV_G_: between-subject biological variation; CV_A_: analytical variation; CI: confidence interval; Aβ: amyloid-β; GFAP: glial fibrillary acidic protein; NfL: neurofilament light; P-tau: phosphorylated tau.

**Table 3.**
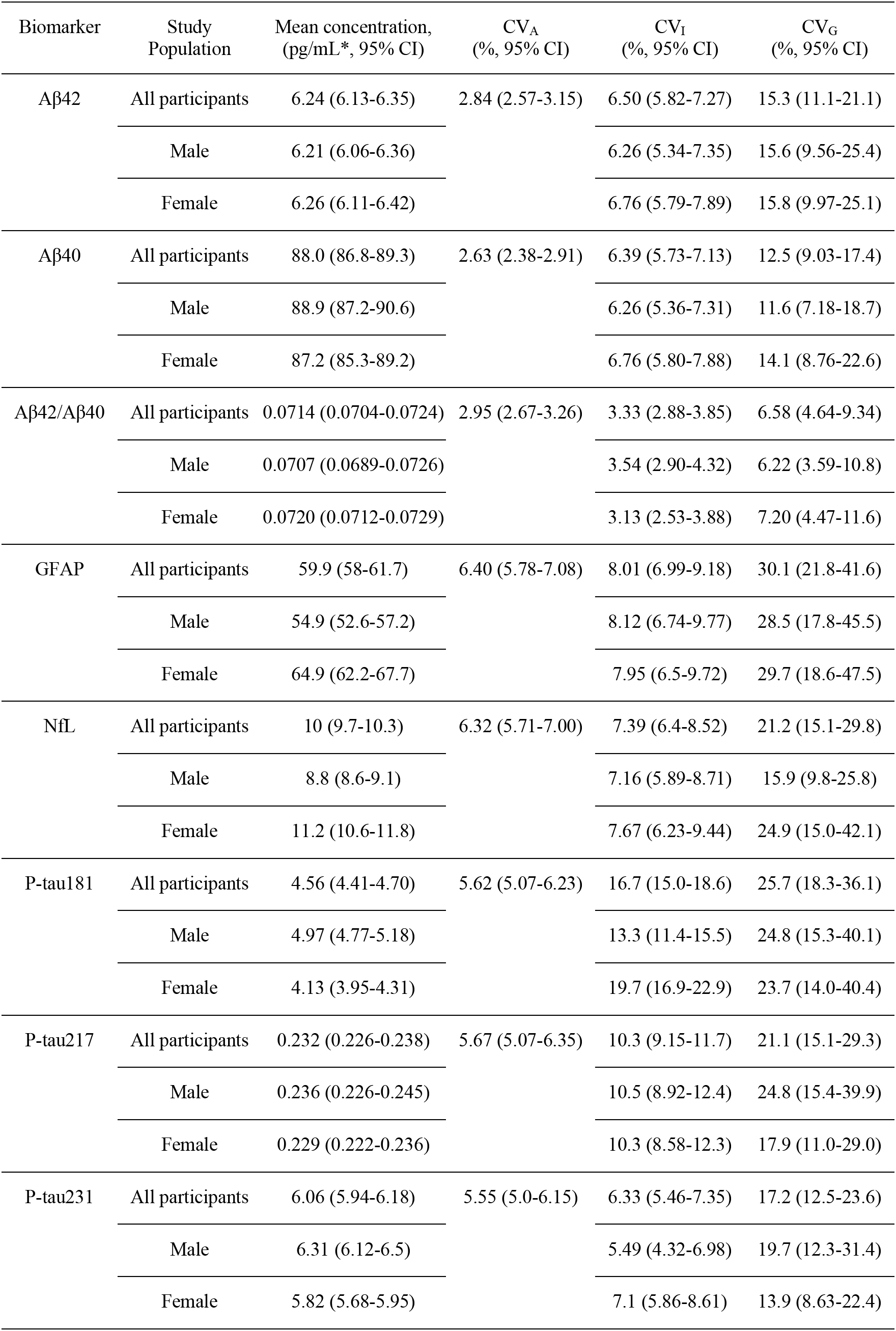
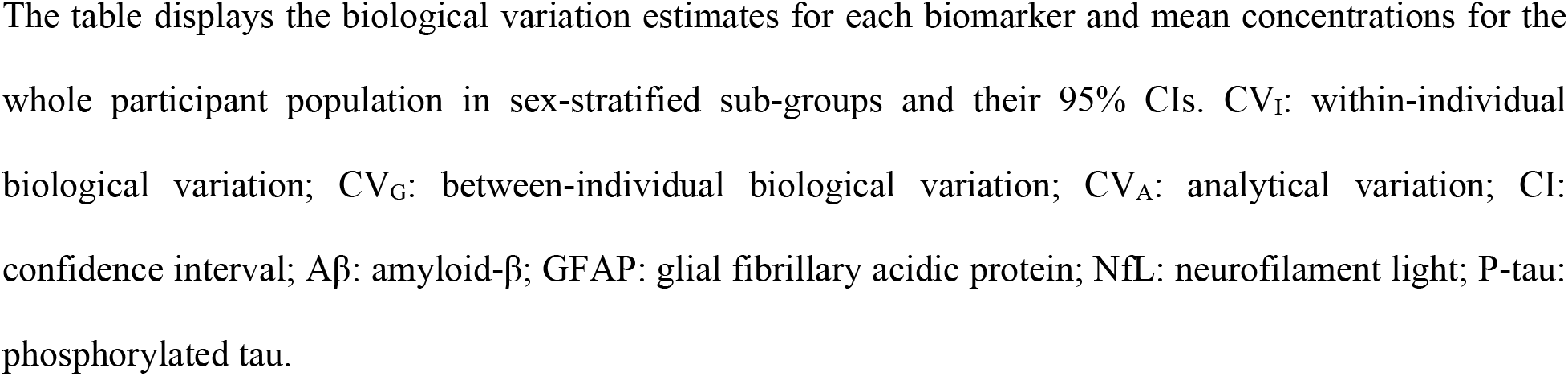
Biological variation estimates for the whole study population and according to sex.

| Biomarker | Study Population | Mean concentration, (pg/mL *, 95% CI) | CV<sub>A</sub> (% , 95% CI) | CV<sub>I</sub> (% , 95% CI) | CV<sub>G</sub> (% , 95% CI) |
| --- | --- | --- | --- | --- | --- |
| Aβ42 | All participants | 6.24 (6.13-6.35) | 2.84 (2.57-3.15) | 6.50 (5.82-7.27) | 15.3 (11.1-21.1) |
|  | Male | 6.21 (6.06-6.36) |  | 6.26 (5.34-7.35) | 15.6 (9.56-25.4) |
|  | Female | 6.26 (6.11-6.42) |  | 6.76 (5.79-7.89) | 15.8 (9.97-25.1) |
| Aβ40 | All participants | 88.0 (86.8-89.3) | 2.63 (2.38-2.91) | 6.39 (5.73-7.13) | 12.5 (9.03-17.4) |
|  | Male | 88.9 (87.2-90.6) |  | 6.26 (5.36-7.31) | 11.6 (7.18-18.7) |
|  | Female | 87.2 (85.3-89.2) |  | 6.76 (5.80-7.88) | 14.1 (8.76-22.6) |
| Aβ42/Aβ40 | All participants | 0.0714 (0.0704-0.0724) | 2.95 (2.67-3.26) | 3.33 (2.88-3.85) | 6.58 (4.64-9.34) |
|  | Male | 0.0707 (0.0689-0.0726) |  | 3.54 (2.90-4.32) | 6.22 (3.59-10.8) |
|  | Female | 0.0720 (0.0712-0.0729) |  | 3.13 (2.53-3.88) | 7.20 (4.47-11.6) |
| GFAP | All participants | 59.9 (58-61.7) | 6.40 (5.78-7.08) | 8.01 (6.99-9.18) | 30.1 (21.8-41.6) |
|  | Male | 54.9 (52.6-57.2) |  | 8.12 (6.74-9.77) | 28.5 (17.8-45.5) |
|  | Female | 64.9 (62.2-67.7) |  | 7.95 (6.5-9.72) | 29.7 (18.6-47.5) |
| NfL | All participants | 10 (9.7-10.3) | 6.32 (5.71-7.00) | 7.39 (6.4-8.52) | 21.2 (15.1-29.8) |
|  | Male | 8.8 (8.6-9.1) |  | 7.16 (5.89-8.71) | 15.9 (9.8-25.8) |
|  | Female | 11.2 (10.6-11.8) |  | 7.67 (6.23-9.44) | 24.9 (15.0-42.1) |
| P-tau181 | All participants | 4.56 (4.41-4.70) | 5.62 (5.07-6.23) | 16.7 (15.0-18.6) | 25.7 (18.3-36.1) |
|  | Male | 4.97 (4.77-5.18) |  | 13.3 (11.4-15.5) | 24.8 (15.3-40.1) |
|  | Female | 4.13 (3.95-4.31) |  | 19.7 (16.9-22.9) | 23.7 (14.0-40.4) |
| P-tau217 | All participants | 0.232 (0.226-0.238) | 5.67 (5.07-6.35) | 10.3 (9.15-11.7) | 21.1 (15.1-29.3) |
|  | Male | 0.236 (0.226-0.245) |  | 10.5 (8.92-12.4) | 24.8 (15.4-39.9) |
|  | Female | 0.229 (0.222-0.236) |  | 10.3 (8.58-12.3) | 17.9 (11.0-29.0) |
| P-tau231 | All participants | 6.06 (5.94-6.18) | 5.55 (5.0-6.15) | 6.33 (5.46-7.35) | 17.2 (12.5-23.6) |
|  | Male | 6.31 (6.12-6.5) |  | 5.49 (4.32-6.98) | 19.7 (12.3-31.4) |
|  | Female | 5.82 (5.68-5.95) |  | 7.1 (5.86-8.61) | 13.9 (8.63-22.4) |

The table displays the biological variation estimates for each biomarker and mean concentrations for the whole participant population in sex-stratified sub-groups and their 95% CIs. $CV_I$ : within-individual biological variation; $CV_G$ : between-individual biological variation; $CV_A$ : analytical variation; CI: confidence interval; $A\beta$ : amyloid-β ; GFAP: glial fibrillary acidic protein; NfL: neurofilament light; P-tau: phosphorylated tau.

### 3.4 Within-subject biological variation (CV_I_)

Figure 2B graphically represents the CV_I_ values and their associated 95% CIs, *i.e.*, how much biomarker concentrations fluctuate around each individual’s homeostatic setpoint. Plasma Aβ42 and Aβ40 demonstrated low and very similar CV_I_’s (Aβ42: 6.5%, 95% CI 5.8-7.3; Aβ40: 6.4%, 95% CI 5.7-7.1), and the plasma Aβ42/Aβ40 ratio demonstrated the lowest CV_I_ among all evaluated biomarkers (3.3%, 95% CI 2.9-3.9). Among plasma p-tau variants, p-tau231 demonstrated the lowest CV_I_ (6.3%, 95% CI 5.5-7.4), followed by p-tau217 (10.3%, 95% CI 9.2-11.7), and by p-tau181 with a considerably higher CV_I_ (16.7%, 95%CI 15.0-18.6). Plasma GFAP also demonstrated a relatively low CV_I_ (8.0%, 95% CI 7.0-9.2), comparable to that observed for NfL (7.4%, 95%CI 6.4-8.5). In Table 3, the CV_I_’s are also shown separately for males and females, an important and needed subgroup analyses in BV studies. Except for p-tau181, no differences in CV_I_ were observed for the evaluated biomarkers, with overlapping 95% CIs for male and female CV_I_’s. For plasma p-tau181, females (19.7%, 95% CI 16.9-22.9) demonstrated a higher CV_I_ than males (13.3%, 95% CI 11.4-15.5).

### 3.5 Between-subject biological variation (CV_G_)

Figure 2C graphically represents the CV_G_ values and their associated 95% CIs, *i.e.*, how much biomarker levels vary between healthy individuals. Among Aβ biomarkers, plasma Aβ42/Aβ40 demonstrated the lowest CV_G_ (6.6%, 95% CI 4.6-9.3), with higher and similar estimates for Aβ42 (15.3%, 95% CI 11.1-21.1) and Aβ40 (12.5%, 95% CI 9.0-17.4). For the other biomarkers, CV_G_’s were generally higher than those for Aβ biomarkers. GFAP demonstrated the highest CV_G_ among all biomarkers (30.1%, 95% CI 21.8-41.6), and slightly higher than that of NfL (21.2%, 95% CI 15.1-29.8). Among p-tau biomarkers, p-tau231 (17.2%, 95% CI 17.2-19.7) demonstrated the lowest CV_G_, followed by p-tau217 (21.1%, 95%CI 15.1-29.3) and p-tau181 (25.7%, 18.3-36.1%). Table 3 indicates the CV_G_’s separately for males and females. No differences in CV_G_ estimates were found for the evaluated biomarkers, with overlapping 95% CIs between males and females for all measurands. Table 3 also shows the mean concentrations and their 95% CIs for males and females separately, with slightly higher concentrations in females observed for plasma GFAP and NfL, and slightly higher concentrations in males for plasma p-tau181 and p-tau231.

### 3.6 Analytical performance specifications and other metrics

Table 4 shows analytical performance specifications (APS) based on the desirable criteria (intermediate stringency), for imprecision, bias. In terms of desirable assay imprecision, the highest demand was for plasma Aβ42/Aβ40 (CV_APS_<1.7%), with the lowest demands for plasma p-tau217 (CV_APS_<5.2%) and p-tau181 (CV_APS_<8.4%). Table 4 shows the estimated reference change values (RCV), as well as the number of samples needed to estimate the homeostatic point. Plasma Aβ42/Aβ40 demonstrated the lowest RCVs needed for a significant decrease (11%) and for na increase (13%). Similar RCVs for both decrease (-20 to -28%) and increase (26 to 38%) were observed for GFAP, NfL, p-tau217 and p-tau231, with the highest RCVs for p-tau181 (decrease: -38.3%; increase: 62.2%).

**Table 4.**
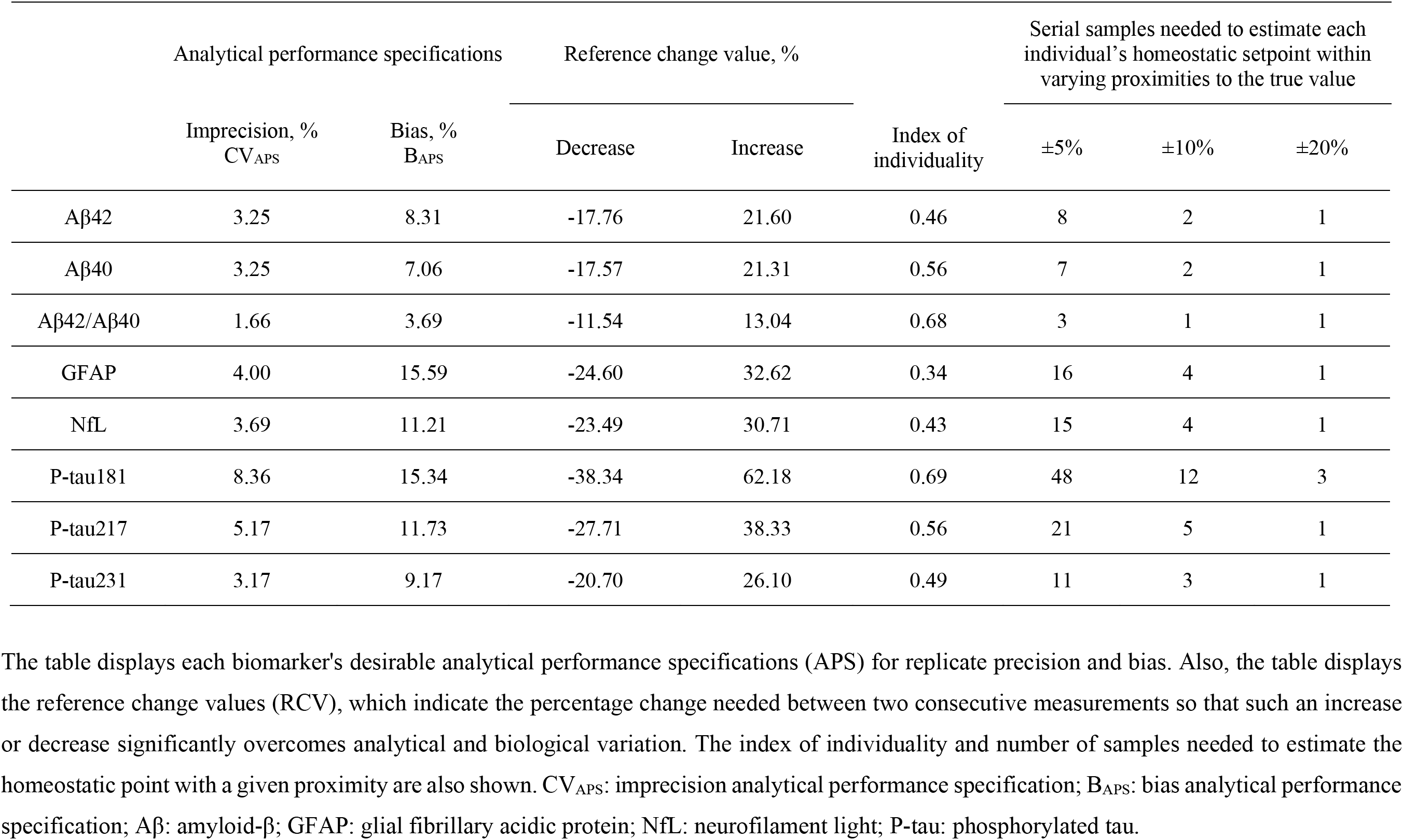
Metrics derived from BV estimates. The table displays each biomarker’s desirable analytical performance specifications (APS) for replicate precision and bias. Also, the table displays the reference change values (RCV), which indicate the percentage change needed between two consecutive measurements so that such an increase or decrease significantly overcomes analytical and biological variation. The index of individuality and number of samples needed to estimate the homeostatic point with a given proximity are also shown. CV_APS_: imprecision analytical performance specification; B_APS_: bias analytical performance specification; Aβ: amyloid-β; GFAP: glial fibrillary acidic protein; NfL: neurofilament light; P-tau: phosphorylated tau.

|  | Analytical performance specifications |  | Reference change value, % |  | Index of individuality | Serial samples needed to estimate each individual's homeostatic setpoint within varying proximities to the true value |  |  |
| --- | --- | --- | --- | --- | --- | --- | --- | --- |
|  | Imprecision, %<br>CV<sub>APS</sub> | Bias, %<br>B<sub>APS</sub> | Decrease | Increase |  | ±5% | ±10% | ±20% |
| Aβ42 | 3.25 | 8.31 | -17.76 | 21.60 | 0.46 | 8 | 2 | 1 |
| Aβ40 | 3.25 | 7.06 | -17.57 | 21.31 | 0.56 | 7 | 2 | 1 |
| Aβ42/Aβ40 | 1.66 | 3.69 | -11.54 | 13.04 | 0.68 | 3 | 1 | 1 |
| GFAP | 4.00 | 15.59 | -24.60 | 32.62 | 0.34 | 16 | 4 | 1 |
| NfL | 3.69 | 11.21 | -23.49 | 30.71 | 0.43 | 15 | 4 | 1 |
| P-tau181 | 8.36 | 15.34 | -38.34 | 62.18 | 0.69 | 48 | 12 | 3 |
| P-tau217 | 5.17 | 11.73 | -27.71 | 38.33 | 0.56 | 21 | 5 | 1 |
| P-tau231 | 3.17 | 9.17 | -20.70 | 26.10 | 0.49 | 11 | 3 | 1 |

## 4. Discussion

In this study, we report BV estimates for novel AD plasma biomarkers generated based on a high number of weekly samples per individual, with a comprehensive biomarker panel measured within the same participants. These are the first reported BV estimates for blood Aβ42, Aβ40, Aβ42/Aβ40, and p-tau, but also for plasma NfL and GFAP, which had their BV previously evaluated in serum.^55–57^ We found that within- and between-subject biological variation can be considerably different for each AD biomarker class, which may inform and impact biomarker application given each clinical or research context.

In addition to improving clinical and research interpretation of these biomarkers, reliable BV data enables the determination of the APS needed for each biomarker. In short, the assay imprecision (i.e. CV_A_) should be considerably lower than the biomarker’s CV_I_, with the desirable imprecision analytical performance being that CV_A_≤CV_I_/2.^58^ Here, CV_A_’s were slightly higher than desired for most biomarkers. Plasma Aβ42 and Aβ40 (but not Aβ42/Aβ40) and plasma p-tau181 were within the desirable performance, with plasma p-tau217 showing a very close to desirable analytical performance (CV_A_: 5.7%; CV_APS_: ≤5.2%). Nevertheless, it is important to emphasize that BV estimates are not standalone criteria to determine APS for assays, but rather a complementary tool to refine the determination of analytical goals given each analyte’s clinical application context. For instance, the higher-than-desirable observed CV_A_’s are likely not a cause of concern in light of the main clinical applications of NfL and p-tau variants, whereas it might affect more Aβ42/Aβ40, as discussed below.

Plasma p-tau is an AD-specific biomarker envisioned to be implemented as a screening tool to classify patients seeking medical advise for cognitive symptoms into high-, intermediate- and low-risk of having AD pathology.^59,60^ The emergence of plasma p-tau assays has sparkled a debate on potential differences between p-tau phospho-sites.^2^ In our study, CV_A_ was remarkably similar for all three plasma p-tau variants, but they demonstrated considerably different CV_I_’s. Interestingly, p-tau231 demonstrated the lowest CV_I_ (6.3%, 95%CI 5.5-7.4%), followed by p-tau217 (10.3%, 95%CI 9.2-11.7%) and p-tau181 (16.7%, 95%CI 12.5-23.6%). These may suggest differences in release, clearance or transportation of plasma p-tau species. Recent head-to-head comparisons of plasma p-tau variants showed the most promising candidates were increased between 100-360% in the presence of AD pathology.^9,10^ Taking the magnitude of these increases into account, it seems reasonable to state that the currently used p-tau assays demonstrate a satisfactory analytical performance for clinical applications, having their diagnostic ability less vulnerable to biological and analytical variation.

Plasma NfL has been successfully introduced in clinical routine in some laboratories, being useful both in a wide range of neurodegenerative diseases and in acute neurological conditions.^22,24,26,61^ Plasma NfL showed a relatively low CV_I_ (7.4%, 95%CI 6.4-8.5%), and a higher CV_G_ (21.2%, 95%CI 15.1-29.8%). This is in accordance with the ∼10% CV_I_ and CVG’s reported in a previous BV study evaluating serum NfL in a Turkish cohort, with a different analytical platform, but also with 10-week weekly collection.^57^ Our RCVs for NfL (increase: +30.7; decrease: -23.4%) also closely agreed with those in that study (increase: +32.7; decrease: -24.7%). A different study in a Danish cohort reported a substantially lower CV_I_ for serum NfL (Simoa) with non-overlapping CIs (CV_I_=3%, 95%CI 1.2-5.0%), and lower RCVs (increase: +24.3; decrease: -19.5%).^56^ The lower NfL CV_I_ in this cohort could be attributed to the shorter sampling period of three consecutive days, which may lead to lower BV estimates, compared with the 10-week weekly collection in both ours and the Turkish study.^62^ The relatively high CV_G_ seen for plasma NfL reflects higher inter-individual variability, posing a challenge to its clinical interpretation in conditions with modest NfL fold-changes such as AD, where the diagnostic utility of NfL is limited. However, in certain clinical scenarios, this between-subject variation is likely of limited importance, including differentiating primary psychiatric disorders from frontotemporal dementia,^63^ or in prognostic evaluation of acute conditions such as cardiac arrest, stroke and traumatic brain injury.^24,26,64^ In these cases, the magnitude of NfL increases is much larger compared to relatively higher CV_G_’s.

We also provide BV estimates for plasma GFAP. While it is not yet clear what GFAP in the blood reflects, with differences against CSF GFAP,^65^ it has been associated with Aβ pathology and demonstrated promising diagnostic performance^18,20^, but with some studies reporting increased levels in Aβ-negative neurodegenerative disorders, e.g. frontotemporal lobar degeneration.^66^ We observed a relatively low CV_I_ for plasma GFAP (8.0%, 95%CI 7.0-9.2), and the highest CV_G_ among all evaluated biomarkers (30.1%, 95%CI 21.8-41.6%). The estimated CV_I_ was similar to a previously reported CV_I_ for serum GFAP (9.7%, 95%CI 7.6-11.8), and while their CV_G_ was slightly higher (39.5%, 95%CI 31.7-47.3), it was still in a similar higher tier, with RCVs also agreeing between studies. Plasma GFAP presents a high CV_G_, which, alongside its poorly understood clinical meaning, indicates potential difficulties for its individual-level interpretation. We report here the first BV estimates for plasma Aβ. Plasma Aβ42/Aβ40 is a widely investigated biomarker associated with Aβ pathology currently in clinical use in to support AD diagnosis.^12,67,68^ However, in Aβ-positive vs Aβ-negative individuals, plasma Aβ42/Aβ40 is only decreased by 8-14%.^13^ This clinical context places, per se, a robustness issue for this biomarker, since the modest disease-related fold-changes are in a similar magnitude to that of common analytical variation results seen in clinical chemistry.^2,14–16^ Our novel BV findings further support that the implementation of plasma Aβ42/Aβ40 will likely face difficulties if introduced in clinical practice. Plasma Aβ42/Aβ40 demonstrated a considerably low CV_I_ (3.3%, 95%CI 2.9-3.9), and a relatively low CV_G_ (6.6%, 95%CI 4.6-9.3). The low CV_I_ introduces a very high demand on analytical performance, since, desirably, the CV_A_ should be less than half of the CV_I_ (CV_APS_=1.7%; Table 4), and optimally, less or equal to a quarter of CV_I_ (CV_I_/4=0.7%). This occurs in a different clinical context to that of plasma p-tau, where larger disease-related increases make the ∼5.5% CV_A_’s acceptable. Additionally, while there are several different plasma Aβ assays currently in use, they present similarity in their small disease-changes.^13^ Considering previous BV studies showing different assay versions for the same analyte present indistinguishable CV_I_’s, it is expected that the CV_I_ for plasma Aβ42/Aβ40 would be similar across assays.^47,52^ Further, the desirable bias is also considerably low for plasma Aβ42/Aβ40 (B_APS_=3.7%; Table 4), and it is unlikely that batch-to-batch variations could be kept low enough to meet it.

Here, we report RCVs for AD plasma biomarkers, a potentially clinically valuable tool not yet explored in the AD field. By enumerating the change that can be explained by biological and analytical variation, when monitoring an individual with consecutive tests over time, more informed clinical decisions can be made. This becomes especially relevant with novel anti-Aβ immunotherapies such as lecanemab and donanemab.^27,69^ These drugs have been shown to substantially reduce plasma p-tau217, in a group-level, as early as at 12 weeks of treatment (with our RCVs derived in a similar time frame), during the inital phase of Aβ-plaque reduction.^27^ RCVs could be potentially used to identify whether a reduction in plasma p-tau217 after treatment initiation could indeed be related to a positive treatment response, provided trial data support an individual-level association between p-tau reduction and benefit. On the other hand, this class of drugs can cause amyloid-related imaging abnormalities (ARIA) of the hemorrhage or edema type, which can be very harmful.^70^ If NfL proves capable of tracking such changes (currently unknown from current trials), RCVs could also be potentially useful to monitor their emergence. However, it is important to take into account that our RCVs were obtained using the presumably healthy cognitively unimpaired sample and assays herein described, and, for this reason, cannot be considered as universal values, and each laboratory has to determine their own RCVs based on their CV_A_ estimates.

Our study provides important novel data for interpreting of AD blood biomarkers. Most published studies have collected samples either cross-sectionally or over longer periods of time (e.g. 6 months, yearly), and little is known about their shorter-term variability, with a BV study like ours providing a unique opportunity to evaluate shorter-term fluctuations. Figure 3 shows plasma p-tau181 and p-tau217 over the study period male subjects with similar ages. Subject “A” shows minimal fluctuation around the homeostatic point, while subject “B” experiences two spikes in plasma p-tau. These fluctuations were not analytical outliers, and since a chronic disease like AD is unlikely to manifest an oscillatory progression from week-to-week in middle-aged adults, there might be yet uncharacterized factors influencing biomarker readings. Clinical decisions made on a single sample collected on a p-tau “spike” day could erroneously classify patients as “abnormal”, as exemplified by two previously described cut-offs.^43,71^ Such high-value outliers are not uncommon in Aβ-negative groups when examining data points from recent cross-sectional studies,^4–6^ and we recommend caution for researchers and clinicians when interpreting AD blood biomaker results from a single sample.

**Figure 3.**
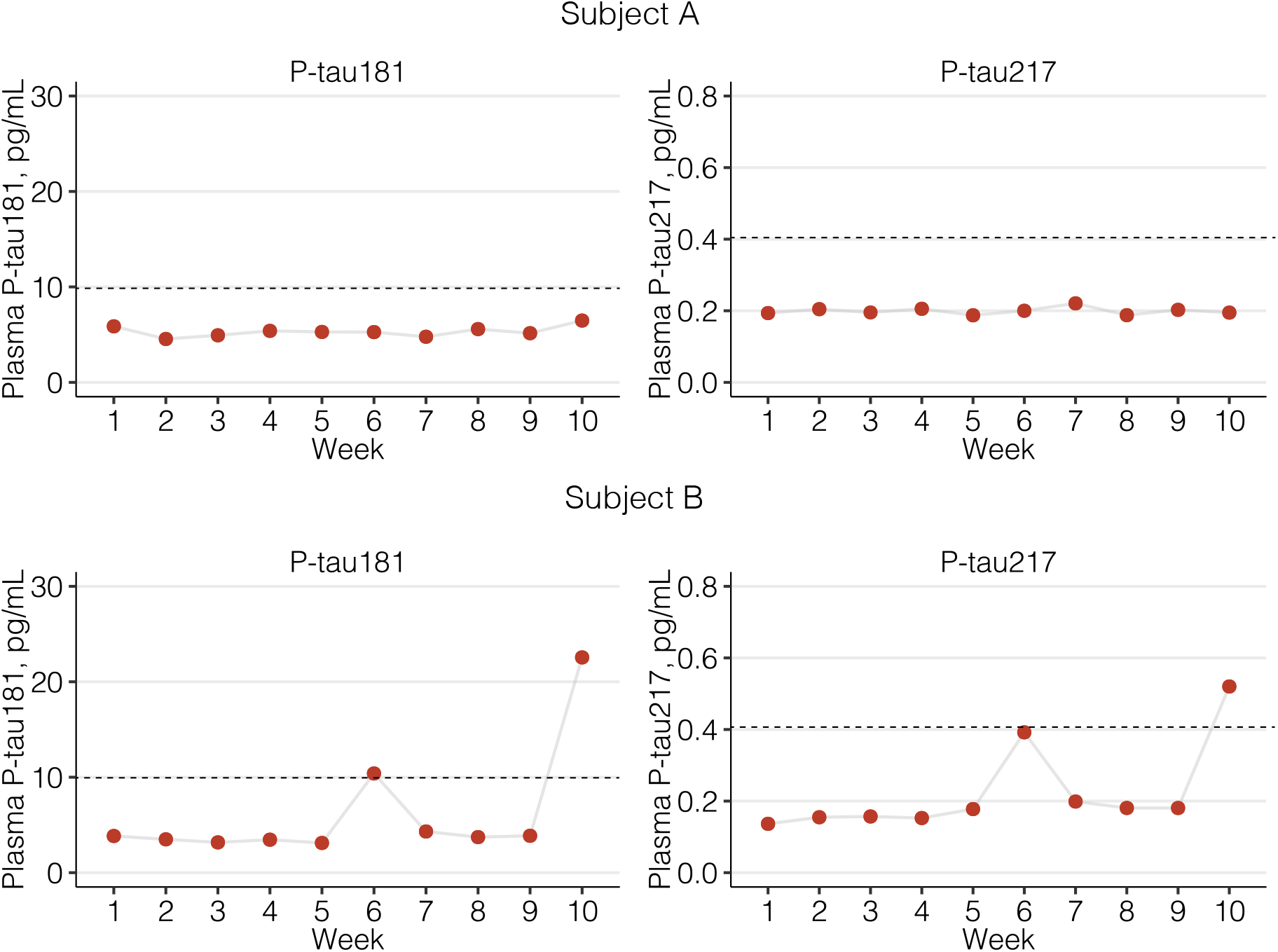
Example of 10-week variability in plasma p-tau181 and p-tau217 in two male study participants in their 40s. The figure shows the variability in plasma p-tau181 and p-tau217 levels over 10 weeks in two study male healthy study participants in their 40s. Dots correspond to the mean concentration of the two duplicate measurements, and all of these subjects’ measurements demonstrated acceptable agreement between replicates, indicating that any deviation observed does not come from analytical imprecision. Dashed lines represent previously published cut-offs for plasma p-tau181 and p-tau217 as illustrative examples of the potential impacts of these fluctuations over decision making. Of note, the outlier data points were excluded from CV_I_ calculations in the homogeneity analyses since they do not reflect the expected homeostatic fluctuation. It is likely that these outlier data points instead correspond to a yet unknown factor that affects biomarker readings. CV_I_: within-individual biological variation; P-tau: phosphorylated tau.

Additionally, knowledge of CV_I_ enables calculating the number of samples needed to estimate the individual’s homeostatic point (HSP) within a certain proximityof the true value. To estimate the true HSPs of all analytes with a deviation of ±20% (α<0.05), a reasonable margin for most analytes here (given clinical contexts discussed above), one sample suffices for all biomarkers but p-tau181 (NHSP=3). Reducing this deviation to 5%, likely needed for plasma Aβ42/Aβ40 (but not for other biomarkers), three samples would be required (Table 4). The II evaluates the utility of reference intervals (RIs). For analytes with pronounced individuality and a relatively low CV_I_ compared to CV_G_ (II<0.6), RCVs are more useful than RIs for accurate interpretation of sequential results, with each individual serving as the optimal reference point for assessing serial. However, RIs remain suitable for analytes with high II (particularly when II>1.4).^33,36^ Here, all II values were below 0.6, except for p-tau181 and Aβ42/Aβ40, which had slightly higher values, indicating marked individuality for these analytes and suggesting the potential for personalized reference intervals.^33,36,72^

This study has limitations. Although well-powered in individual-level serial sampling, the number of participants was relatively small, possibly affecting CV_G_ more than CV_I_ estimates. We found some concentration differences between males and females, and an unexpected sex difference in CV_I_ for p-tau181, warranting further studies. The relatively younger population studied here may not capture biomarker fluctuations related to factors such as co-morbidities and medication use.^73^ Lack of confirmatory CSF or PET biomarkers prevented us from evaluating the effects of AD pathology over BV estimates. Further evaluation on more diverse populations is also needed. Our study also has a number of strengths, involving quantifying a comprehensive panel of AD biomarkers in a dataset following all EFLM recommendations for BV studies,^30,31^ that has generated reliable BV data for many other analytes.^37,38^ In summary, our study provides novel information on the BV of novel AD plasma biomarkers, with important considerations for research, trials and clinical practice.

## Data Availability

All data produced in the present study are available upon reasonable request to the authors

## Acknowledgements

HZ is a Wallenberg Scholar supported by grants from the Swedish Research Council (#2022-01018 and #2019-02397), the European Union’s Horizon Europe research and innovation programme under grant agreement No 101053962, Swedish State Support for Clinical Research (#ALFGBG-71320), the Alzheimer Drug Discovery Foundation (ADDF), USA (#201809-2016862), the AD Strategic Fund and the Alzheimer’s Association (#ADSF-21-831376-C, #ADSF-21-831381-C, and #ADSF-21-831377-C), the Bluefield Project, the Olav Thon Foundation, the Erling-Persson Family Foundation, Stiftelsen för Gamla Tjänarinnor, Hjärnfonden, Sweden (#FO2022-0270), the European Union’s Horizon 2020 research and innovation programme under the Marie Skłodowska-Curie grant agreement No 860197 (MIRIADE), the European Union Joint Programme – Neurodegenerative Disease Research (JPND2021-00694), the National Institute for Health and Care Research University College London Hospitals Biomedical Research Centre, and the UK Dementia Research Institute at UCL (UKDRI-1003). TKK was supported by the NIH (P30 AG066468, RF1 AG052525-01A1, R01 AG053952-05, R37 AG023651-17, RF1 AG025516-12A1, R01 AG073267-02, R01 AG075336-01, R01 AG072641-02, P01 AG025204-16), the Swedish Research Council (Vetenskåpradet; #2021-03244), the Alzheimer’s Association (#AARF-21-850325), the Swedish Alzheimer Foundation (Alzheimerfonden), the Aina (Ann) Wallströms and Mary-Ann Sjöbloms stiftelsen, and the Emil och Wera Cornells stiftelsen. KB is supported by the Swedish Research Council (#2017-00915 and #2022-00732), the Swedish Alzheimer Foundation (#AF-930351, #AF-939721 and #AF-968270), Hjärnfonden, Sweden (#FO2017-0243 and #ALZ2022-0006), the Swedish state under the agreement between the Swedish government and the County Councils, the ALF-agreement (#ALFGBG-715986 and #ALFGBG-965240), the European Union Joint Program for Neurodegenerative Disorders (JPND2019-466-236), the Alzheimer’s Association 2021 Zenith Award (ZEN-21-848495), and the Alzheimer’s Association 2022-2025 Grant (SG-23-1038904 QC).

## References

1. Hansson O. Biomarkers for neurodegenerative diseases. Nat Med. 2021;27(6):954–963. doi:10.1038/s41591-021-01382-x

2. Karikari TK, Ashton NJ, Brinkmalm G, et al. Blood phospho-tau in Alzheimer disease: analysis, interpretation, and clinical utility. Nat Rev Neurol. 2022;18(7):400–418. doi:10.1038/s41582-022-00665-2

3. Jack CR, Bennett DA, Blennow K, et al. NIA-AA Research Framework: Toward a biological definition of Alzheimer’s disease. Alzheimers Dement. 2018;14(4):535–562. doi:10.1016/j.jalz.2018.02.018

4. Palmqvist S, Janelidze S, Quiroz YT, et al. Discriminative Accuracy of Plasma Phospho-tau217 for Alzheimer Disease vs Other Neurodegenerative Disorders. JAMA. 2020;324(8):772–781. doi:10.1001/jama.2020.12134

5. Karikari TK, Pascoal TA, Ashton NJ, et al. Blood phosphorylated tau 181 as a biomarker for Alzheimer’s disease: a diagnostic performance and prediction modelling study using data from four prospective cohorts. Lancet Neurol. 2020;19(5):422–433. doi:10.1016/S1474-4422(20)30071-5

6. Ashton NJ, Pascoal TA, Karikari TK, et al. Plasma p-tau231: a new biomarker for incipient Alzheimer’s disease pathology. Acta Neuropathol. 2021;141(5):709–724. doi:10.1007/s00401-021-02275-6

7. Ashton NJ, Janelidze S, Mattsson-Carlgren N, et al. Differential roles of Aβ42/40, p-tau231 and p-tau217 for Alzheimer’s trial selection and disease monitoring. Nat Med. Published online December 1, 2022:1–8. doi:10.1038/s41591-022-02074-w

8. Montoliu-Gaya L, Benedet AL, Tissot C, et al. Mass spectrometric simultaneous quantification of tau species in plasma shows differential associations with amyloid and tau pathologies. Nat Aging. Published online April 27, 2023:1–9. doi:10.1038/s43587-023-00405-1

9. Janelidze S, Bali D, Ashton NJ, et al. Head-to-head comparison of 10 plasma phospho-tau assays in prodromal Alzheimer’s disease. Brain. Published online September 10, 2022:awac333. doi:10.1093/brain/awac333

10. Ashton NJ, Puig-Pijoan A, Milà-Alomà M, et al. Plasma and CSF biomarkers in a memory clinic: Head-to-head comparison of phosphorylated tau immunoassays. Alzheimer’s & Dementia. n/a(n/a). doi:10.1002/alz.12841

11. Milà-Alomà M, Ashton NJ, Shekari M, et al. Plasma p-tau231 and p-tau217 as state markers of amyloid-β pathology in preclinical Alzheimer’s disease. Nat Med. 2022;28(9):1797–1801. doi:10.1038/s41591-022-01925-w

12. Schindler SE, Bollinger JG, Ovod V, et al. High-precision plasma β-amyloid 42/40 predicts current and future brain amyloidosis. Neurology. 2019;93(17):e1647–e1659. doi:10.1212/WNL.0000000000008081

13. Janelidze S, Teunissen CE, Zetterberg H, et al. Head-to-Head Comparison of 8 Plasma Amyloid-β 42/40 Assays in Alzheimer Disease. JAMA Neurol. 2021;78(11):1375–1382. doi:10.1001/jamaneurol.2021.3180

14. Benedet AL, Brum WS, Hansson O, et al. The accuracy and robustness of plasma biomarker models for amyloid PET positivity. Alzheimers Res Ther. 2022;14(1):26. doi:10.1186/s13195-021-00942-0

15. Rabe C, Bittner T, Jethwa A, et al. Clinical performance and robustness evaluation of plasma amyloid-β42/40 prescreening. Alzheimers Dement. 2023;19(4):1393–1402. doi:10.1002/alz.12801

16. Cullen NC, Janelidze S, Mattsson-Carlgren N, et al. Test-retest variability of plasma biomarkers in Alzheimer’s disease and its effects on clinical prediction models. Alzheimers Dement. Published online June 14, 2022. doi:10.1002/alz.12706

17. Pekny M, Nilsson M. Astrocyte activation and reactive gliosis. Glia. 2005;50(4):427–434. doi:10.1002/glia.20207

18. Benedet AL, Milà-Alomà M, Vrillon A, et al. Differences Between Plasma and Cerebrospinal Fluid Glial Fibrillary Acidic Protein Levels Across the Alzheimer Disease Continuum. JAMA Neurology. 2021;78(12):1471–1483. doi:10.1001/jamaneurol.2021.3671

19. Pereira JB, Janelidze S, Smith R, et al. Plasma GFAP is an early marker of amyloid-β but not tau pathology in Alzheimer’s disease. Brain. 2021;144(11):3505–3516. doi:10.1093/brain/awab223

20. Shen XN, Huang SY, Cui M, et al. Plasma Glial Fibrillary Acidic Protein in the Alzheimer Disease Continuum: Relationship to Other Biomarkers, Differential Diagnosis, and Prediction of Clinical Progression. Clin Chem. Published online March 2, 2023:hvad018. doi:10.1093/clinchem/hvad018

21. Montoliu-Gaya L, Alcolea D, Ashton NJ, et al. Plasma and cerebrospinal fluid glial fibrillary acidic protein levels in adults with Down syndrome: a longitudinal cohort study. EBioMedicine. 2023;90:104547. doi:10.1016/j.ebiom.2023.104547

22. Ashton NJ, Janelidze S, Al Khleifat A, et al. A multicentre validation study of the diagnostic value of plasma neurofilament light. Nat Commun. 2021;12(1):3400. doi:10.1038/s41467-021-23620-z

23. Mattsson N, Cullen NC, Andreasson U, Zetterberg H, Blennow K. Association Between Longitudinal Plasma Neurofilament Light and Neurodegeneration in Patients With Alzheimer Disease. Jama Neurol. 2019;76(7). doi:10.1001/jamaneurol.2019.0765

24. Gendron TF, Badi MK, Heckman MG, et al. Plasma neurofilament light predicts mortality in patients with stroke. Science Translational Medicine. 2020;12(569):eaay1913. doi:10.1126/scitranslmed.aay1913

25. Michaëlsson I, Hallén T, Carstam L, et al. Circulating Brain Injury Biomarkers: A Novel Method for Quantification of the Impact on the Brain After Tumor Surgery. Neurosurgery. Published online May 5, 2023. doi:10.1227/neu.0000000000002510

26. Ashton NJ, Moseby-Knappe M, Benedet AL, et al. Alzheimer Disease Blood Biomarkers in Patients With Out-of-Hospital Cardiac Arrest. JAMA Neurology. Published online March 6, 2023. doi:10.1001/jamaneurol.2023.0050

27. Pontecorvo MJ, Lu M, Burnham SC, et al. Association of Donanemab Treatment With Exploratory Plasma Biomarkers in Early Symptomatic Alzheimer Disease: A Secondary Analysis of the TRAILBLAZER-ALZ Randomized Clinical Trial. JAMA Neurology. Published online October 17, 2022. doi:10.1001/jamaneurol.2022.3392

28. Hansson O, Edelmayer RM, Boxer AL, et al. The Alzheimer’s Association appropriate use recommendations for blood biomarkers in Alzheimer’s disease. Alzheimers Dement. 2022;18(12):2669–2686. doi:10.1002/alz.12756

29. Fraser GG, Harris EK. Generation and Application of Data on Biological Variation in Clinical Chemistry. Critical Reviews in Clinical Laboratory Sciences. 1989;27(5):409–437. doi:10.3109/10408368909106595

30. Bartlett WA, Braga F, Carobene A, et al. A checklist for critical appraisal of studies of biological variation. Clin Chem Lab Med. 2015;53(6):879–885. doi:10.1515/cclm-2014-1127

31. Aarsand AK, Røraas T, Fernandez-Calle P, et al. The Biological Variation Data Critical Appraisal Checklist: A Standard for Evaluating Studies on Biological Variation. Clin Chem. 2018;64(3):501–514. doi:10.1373/clinchem.2017.281808

32. Fraser CG. Reference change values: the way forward in monitoring. Ann Clin Biochem. 2009;46(Pt 3):264–265. doi:10.1258/acb.2009.009006

33. Coşkun A, Sandberg S, Unsal I, et al. Personalized Reference Intervals in Laboratory Medicine: A New Model Based on Within-Subject Biological Variation. Clin Chem. 2021;67(2):374–384. doi:10.1093/clinchem/hvaa233

34. Fraser CG, Petersen PH. Analytical performance characteristics should be judged against objective quality specifications. Clin Chem. 1999;45(3):321–323.

35. Haeckel R, Wosniok W, Kratochvila J, Carobene A. A pragmatic proposal for permissible limits in external quality assessment schemes with a compromise between biological variation and the state of the art. Clin Chem Lab Med. 2012;50(5):833–839. doi:10.1515/cclm-2011-0862

36. Carobene A, Banfi G, Locatelli M, Vidali M. Within-person biological variation estimates from the European Biological Variation Study (EuBIVAS) for serum potassium and creatinine used to obtain personalized reference intervals. Clin Chim Acta. 2021;523:205–207. doi:10.1016/j.cca.2021.09.018

37. Carobene A, Strollo M, Jonker N, et al. Sample collections from healthy volunteers for biological variation estimates’ update: a new project undertaken by the Working Group on Biological Variation established by the European Federation of Clinical Chemistry and Laboratory Medicine. Clin Chem Lab Med. 2016;54(10):1599–1608. doi:10.1515/cclm-2016-0035

38. Carobene A, Aarsand AK, Bartlett WA, et al. The European Biological Variation Study (EuBIVAS): a summary report. Clin Chem Lab Med. 2022;60(4):505–517. doi:10.1515/cclm-2021-0370

39. Cavalier E, Lukas P, Bottani M, et al. European Biological Variation Study (EuBIVAS): within- and between-subject biological variation estimates of β-isomerized C-terminal telopeptide of type I collagen (β-CTX), N-terminal propeptide of type I collagen (PINP), osteocalcin, intact fibroblast growth factor 23 and uncarboxylated-unphosphorylated matrix-Gla protein-a cooperation between the EFLM Working Group on Biological Variation and the International Osteoporosis Foundation-International Federation of Clinical Chemistry Committee on Bone Metabolism. Osteoporos Int. 2020;31(8):1461–1470. doi:10.1007/s00198-020-05362-8

40. Clouet-Foraison N, Marcovina SM, Guerra E, et al. Analytical Performance Specifications for Lipoprotein(a), Apolipoprotein B-100, and Apolipoprotein A-I Using the Biological Variation Model in the EuBIVAS Population. Clin Chem. 2020;66(5):727–736. doi:10.1093/clinchem/hvaa054

41. Aarsand AK, Kristoffersen AH, Sandberg S, et al. The European Biological Variation Study (EuBIVAS): Biological Variation Data for Coagulation Markers Estimated by a Bayesian Model. Clin Chem. 2021;67(9):1259–1270. doi:10.1093/clinchem/hvab100

42. Bottani M, Aarsand AK, Banfi G, et al. European Biological Variation Study (EuBIVAS): within- and between-subject biological variation estimates for serum thyroid biomarkers based on weekly samplings from 91 healthy participants. Clin Chem Lab Med. 2022;60(4):523–532. doi:10.1515/cclm-2020-1885

43. Ashton NJ, Brum WS, Molfetta G di, et al. Diagnostic accuracy of the plasma ALZpath pTau217 immunoassay to identify Alzheimers disease pathology. Published online July 12, 2023:2023.07.11.23292493. doi:10.1101/2023.07.11.23292493

44. Ashton NJ, Suárez-Calvet M, Karikari TK, et al. Effects of pre-analytical procedures on blood biomarkers for Alzheimer’s pathophysiology, glial activation, and neurodegeneration. Alzheimers Dement (Amst). 2021;13(1):e12168. doi:10.1002/dad2.12168

45. Verberk IMW, Misdorp EO, Koelewijn J, et al. Characterization of pre-analytical sample handling effects on a panel of Alzheimer’s disease–related blood-based biomarkers: Results from the Standardization of Alzheimer’s Blood Biomarkers (SABB) working group. Alzheimer’s & Dementia. 2022;18(8):1484–1497. doi:10.1002/alz.12510

46. Carobene A, Røraas T, Sølvik UØ, et al. Biological Variation Estimates Obtained from 91 Healthy Study Participants for 9 Enzymes in Serum. Clin Chem. 2017;63(6):1141–1150. doi:10.1373/clinchem.2016.269811

47. Carobene A, Marino I, Coşkun A, et al. The EuBIVAS Project: Within- and Between-Subject Biological Variation Data for Serum Creatinine Using Enzymatic and Alkaline Picrate Methods and Implications for Monitoring. Clin Chem. 2017;63(9):1527–1536. doi:10.1373/clinchem.2017.275115

48. Aarsand AK, Díaz-Garzón J, Fernandez-Calle P, et al. The EuBIVAS: Within- and Between-Subject Biological Variation Data for Electrolytes, Lipids, Urea, Uric Acid, Total Protein, Total Bilirubin, Direct Bilirubin, and Glucose. Clin Chem. 2018;64(9):1380–1393. doi:10.1373/clinchem.2018.288415

49. Carobene A, Guerra E, Locatelli M, et al. Providing Correct Estimates of Biological Variation-Not an Easy Task. The Example of S100-β Protein and Neuron-Specific Enolase. Clin Chem. 2018;64(10):1537–1539. doi:10.1373/clinchem.2018.292169

50. Carobene A, Guerra E, Locatelli M, et al. Biological variation estimates for prostate specific antigen from the European Biological Variation Study; consequences for diagnosis and monitoring of prostate cancer. Clinica Chimica Acta. 2018;486:185–191. doi:10.1016/j.cca.2018.07.043

51. Røraas T, Støve B, Petersen PH, Sandberg S. Biological Variation: The Effect of Different Distributions on Estimated Within-Person Variation and Reference Change Values. Clin Chem. 2016;62(5):725–736. doi:10.1373/clinchem.2015.252296

52. Ceriotti F, Díaz-Garzón Marco J, Fernández-Calle P, et al. The European Biological Variation Study (EuBIVAS): weekly biological variation of cardiac troponin I estimated by the use of two different high-sensitivity cardiac troponin I assays. Clin Chem Lab Med. 2020;58(10):1741–1747. doi:10.1515/cclm-2019-1182

53. Røraas T, Støve B, Petersen PH, Sandberg S. Biological variation: Evaluation of methods for constructing confidence intervals for estimates of within-person biological variation for different distributions of the within-person effect. Clinica Chimica Acta. 2017;468:166–173. doi:10.1016/j.cca.2017.02.021

54. Graybill RKB Franklin A. Confidence Intervals on Variance Components. CRC Press; 2014. doi:10.1201/9781482277142

55. Christensen SH, Hviid CVB, Madsen AT, Parkner T, Winther-Larsen A. Short-term biological variation of serum glial fibrillary acidic protein. Clinical Chemistry and Laboratory Medicine (CCLM). 2022;60(11):1813–1819. doi:10.1515/cclm-2022-0480

56. Hviid CVB, Madsen AT, Winther-Larsen A. Biological variation of serum neurofilament light chain. Clinical Chemistry and Laboratory Medicine (CCLM). 2022;60(4):569–575. doi:10.1515/cclm-2020-1276

57. Carobene A, Maiese K, Aboud-Diwan C, Serteser M, Coskun A, Unsal I. Biological variation estimates for serum neurofilament light chain in healthy subjects. Under review. Published online 2023.

58. Sandberg S, Fraser CG, Horvath AR, et al. Defining analytical performance specifications: Consensus Statement from the 1st Strategic Conference of the European Federation of Clinical Chemistry and Laboratory Medicine. Clinical Chemistry and Laboratory Medicine (CCLM). 2015;53(6):833–835. doi:10.1515/cclm-2015-0067

59. Brum WS, Cullen NC, Janelidze S, et al. A two-step workflow based on plasma p-tau217 to screen for Aβ-positivity with further confirmatory testing only in uncertain cases. Nature Aging. In press. doi:10.1038/s43587-023-00471-5

60. Hansson O, Blennow K, Zetterberg H, Dage J. Blood biomarkers for Alzheimer’s disease in clinical practice and trials. Nat Aging. 2023;3(5):506–519. doi:10.1038/s43587-023-00403-3

61. Simrén J, Andreasson U, Gobom J, et al. Establishment of reference values for plasma neurofilament light based on healthy individuals aged 5–90 years. Brain Communications. 2022;4(4):fcac174. doi:10.1093/braincomms/fcac174

62. Coskun A, Braga F, Carobene A, et al. Systematic review and meta-analysis of within-subject and between-subject biological variation estimates of 20 haematological parameters. Clin Chem Lab Med. 2019;58(1):25–32. doi:10.1515/cclm-2019-0658

63. Al Shweiki MR, Steinacker P, Oeckl P, et al. Neurofilament light chain as a blood biomarker to differentiate psychiatric disorders from behavioural variant frontotemporal dementia. J Psychiatr Res. 2019;113:137–140. doi:10.1016/j.jpsychires.2019.03.019

64. Shahim P, Politis A, Merwe A van der, et al. Neurofilament light as a biomarker in traumatic brain injury. Neurology. 2020;95(6):e610–e622. doi:10.1212/WNL.0000000000009983

65. Simrén J, Weninger H, Brum WS, et al. Differences between blood and cerebrospinal fluid glial fibrillary Acidic protein levels: The effect of sample stability. Alzheimers Dement. 2022;18(10):1988–1992. doi:10.1002/alz.12806

66. Benussi A, Cantoni V, Rivolta J, et al. Classification accuracy of blood-based and neurophysiological markers in the differential diagnosis of Alzheimer’s disease and frontotemporal lobar degeneration. Alzheimers Res Ther. 2022;14(1):155. doi:10.1186/s13195-022-01094-5

67. Nakamura A, Kaneko N, Villemagne VL, et al. High performance plasma amyloid-β biomarkers for Alzheimer’s disease. Nature. 2018;554(7691):249-254. doi:10.1038/nature25456

68. Monane M, Johnson KG, Snider BJ, et al. A blood biomarker test for brain amyloid impacts the clinical evaluation of cognitive impairment. Ann Clin Transl Neurol. Published online August 7, 2023. doi:10.1002/acn3.51863

69. van Dyck CH, Swanson CJ, Aisen P, et al. Lecanemab in Early Alzheimer’s Disease. New England Journal of Medicine. 2022;0(0):null. doi:10.1056/NEJMoa2212948

70. Villain N, Planche V, Levy R. High-clearance anti-amyloid immunotherapies in Alzheimer’s disease. Part 1: Meta-analysis and review of efficacy and safety data, and medico-economical aspects. Revue Neurologique. 2022;178(10):1011–1030. doi:10.1016/j.neurol.2022.06.012

71. Clark C, Lewczuk P, Kornhuber J, et al. Plasma neurofilament light and phosphorylated tau 181 as biomarkers of Alzheimer’s disease pathology and clinical disease progression. Alzheimer’s Research & Therapy. 2021;13(1):65. doi:10.1186/s13195-021-00805-8

72. Carobene A, Banfi G, Locatelli M, Vidali M. Personalized reference intervals: From the statistical significance to the clinical usefulness. Clin Chim Acta. 2022;524:203–204. doi:10.1016/j.cca.2021.10.036

73. Mielke MM, Dage JL, Frank RD, et al. Performance of plasma phosphorylated tau 181 and 217 in the community. Nat Med. 2022;28(7):1398–1405. doi:10.1038/s41591-022-01822-2

